# Exploring the effects of prescribed fire on ticks spread and propagation in a spatial setting

**DOI:** 10.1101/2022.01.12.22268825

**Authors:** Alexander Fulk, Weizhang Huang, Folashade Agusto

## Abstract

Lyme disease is one of the most prominent tick-borne diseases in the United States and prevalence of the disease has been steadily increasing over the past several decades due to a number of factors, including climate change. Methods for control of the disease have been considered, one of which is prescribed burning. In this paper the effects of prescribed burns on the abundance of ticks present in a spatial domain are assessed. A spatial stage-structured tick-host model with an impulsive differential equation system is developed to simulate the effect that controlled burning has on tick populations. Subsequently, a global sensitivity analysis is performed to evaluate the effect of various model parameters on the prevalence of infectious nymphs. Results indicate that while ticks can recover relatively quickly following a burn, yearly, high-intensity prescribed burns can reduce the prevalence of ticks in and around the area that is burned. The use of prescribed burns in preventing the establishment of ticks into new areas is also explored and it is observed that frequent burning can slow establishment considerably.

## 1 Introduction

Since its discovery in the United States in the 1970s, Lyme disease has become one of the most prevalent tick-borne diseases therein [1] while the prevalence of tick-borne diseases in general has been increasing around the world [2]. Lyme disease is one of the most debilitating tick-borne diseases if left untreated [3]. It spreads via a diseased tick which carries and transmits the bacterium, *Borellia Burgdorferi*. Several *Ixodes* tick species can transmit the bacterium that causes Lyme disease, including *I. pacificus, I. ricinus, I. persulcatus*, and *I. scapularis. Ixodes scapularis*, also known as the black-legged tick, is one of the primary vectors for Lyme disease. These ticks can both carry and transmit *B. Burgdorferi* and keep Lyme disease endemic in many parts of the world when combined with their hosts [2, 4, 5]. *B. Burgdorferi* spreads mostly via ticks and their hosts, which is significant because ticks are generalists, meaning that they feed on many hosts, basically whichever hosts are available in a given environment. This fact may have also conditioned the bacterium to evolve to be a generalist pathogen as well, which only adds to the difficulty of reduction or eradication of Lyme disease [6].

There are about 30,000 cases of Lyme disease reported each year in the United States and this number has risen steadily for over two decades. The actual prevalence of the disease is estimated to be about 450,000 cases per year [7]. Various factors influence the surveillance of this particular disease. One such factor is the variability in both symptoms reported and severity of those symptoms. The short-term effects of infection vary widely and are generally not very severe. They include rash or lymph nodes, fever, chills, fatigue, joint and muscle pain, and Erythema (EM) or Bull’s Eye rash. Lyme disease is almost never fatal, however, the long-term effects of infection can be very detrimental as they include severe headaches and neck stiffness, an increase in EM rashes, joint swelling (particularly in large joints like the knee and elbow), facial palsy, dizziness, as well as other neurological effects. Due to these wide-ranging symptoms, this disease is also difficult to diagnose and not enough research has been done to develop an effective vaccine for the disease, although two are currently being tested and refined [3]. There is also well-founded fear that the geographic range of the disease is expanding and in addition to that increased public health threat, new tick-borne pathogens could emerge from mutations of the disease [2, 7, 8, 9, 10]. Though this disease is almost never fatal, the excess strain on public healthcare systems, as well as the potential of mutation of the disease into a more virulent strain mean that reduction of cases is of paramount importance.

There are many possible methods of control available for Lyme disease such as education on best practices when entering an area where ticks are known to inhabit, culling of deer, the primary host for adult females, and the use of pesticides [11]. Each of these methods has its own benefits and drawbacks, but one method in particular that has seen rising popularity among researchers in recent decades is prescribed fire [12, 13, 14, 15, 16, 17]. This method can significantly reduce the number of ticks in an area and it can also destroy forest litter that ticks typically dwell in, making reestablishment more difficult. It is relatively cost-effective and easy to implement and in some cases, essential to the environment [18].

Several studies have been conducted that have tried to elucidate both the short and long-term effects of prescribed burns on tick populations [12, 13, 14, 15, 16, 17, 19, 20, 21, 22, 23, 24, 25, 26, 27, 28]. Some studies showed that in the time immediately following a prescribed burn there is a considerable reduction in the number of ticks at the burn site [12, 13, 14, 15, 19, 20, 21, 22, 23, 24, 25]. Some studies have found contradictory evidence on this short-term effect, but those studies largely did not reflect the reality of prescribed burning [13, 14, 15, 17, 20, 22, 24, 28]. Those studies focused on small plots of land and in some cases, used previously unburned sites, which is the opposite of what is typically seen with prescribed burning. Studies on the long-term effects of prescribed burning are few and far between. More recent studies have shown that frequent prescribed burns can effectively reduce the prevalence of ticks and even the prevalence of certain tick-borne diseases [12, 19].

Regardless of whether or not prescribed burns reduce the prevalence of disease in ticks, the benefits of greatly reduced tick populations should not be understated. With a reduction of the prevalence of ticks in an area, there is also a reduction in encounter rates with humans and thus a reduction in the number of humans infected with Lyme disease. This indicates that regions struggling with Lyme disease that are also well-suited for prescribed burns may have a simple, effective method for reducing the annual number of cases of human Lyme disease.

In this study, we develop a mathematical model that incorporates the effect of prescribed burning in a spatially explicit manner in order to investigate both short-tern and long-term effects of burning on tick populations in different scenarios. Our goal is to study the effect of persistent prescribed burns on the prevalence of ticks over time.

The paper is outlined as follows. The mathematical model is formulated in Section 2. In Section 3 the finite element method used for the numerical exploration is described and then a global sensitivity analysis of the model parameters is carried out to identify the parameters with the most impact on the number of infectious nymphs. In Section 4, the effects of prescribed burning and diffusion are examined in different scenarios that may arise in various environments in the real world. The results are discussed in Section 5 and the conclusions are drawn from the results of the numerical simulations and directions for future research are discussed in Section 6.

## 2 Model formulation

The model used in this study is adapted from Guo and Agusto [29] which investigates the effect of fire intensity (low and high intensities) and duration of the burn on tick population and disease prevalence. We extend their model by adding spatial dependencies on the model variables. The variables included in this model are given as follows: eggs *S*_*E*_ = *S*_*E*_(*x, y, t*), susceptible larvae *S*_*L*_ = *S*_*L*_(*x, y, t*), infectious larvae *I*_*L*_ = *I*_*L*_(*x, y, t*), susceptible nymphs *S*_*N*_ = *S*_*N*_ (*x, y, t*), infectious nymphs *I*_*N*_ = *I*_*N*_ (*x, y, t*), susceptible adults *S*_*A*_ = *S*_*A*_(*x, y, t*), infectious adults *I*_*A*_ = *I*_*A*_(*x, y, t*), susceptible mice *S*_*M*_ = *S*_*M*_ (*x, y, t*), and infectious mice *I*_*M*_ = *I*_*M*_ (*x, y, t*). These variables are defined over the two-dimensional rectangle, Ω = (*a, b*) × (*c, d*). Moreover, we add diffusion into this model in order to evaluate the effect of movement of populations on the viability of prescribed fire.

To model the dynamics of tick eggs, we assume that both susceptible and infectious adult female ticks contribute to egg-laying at a rate *π*_*T*_, which is limited by the carrying capacity, *K* [30, 31]. Maturation of eggs into larvae occurs at the rate *σ*_*T*_, while mortality or inviability is given by the rate *μ*_*E*_. There is no diffusion component for eggs since the female often lay eggs in and under leaf litter after dropping off of the hosts following successful feeding. There is some evidence for transovarial transmission of the family of the bacterium that causes Lyme disease among ticks [32], however it seems that *B. Burgdorferi* is not able to be transmitted [33], thus we do not incorporate an infected egg compartment into the model. Thus, the equation governing how the susceptible egg population changes across time and space reads as

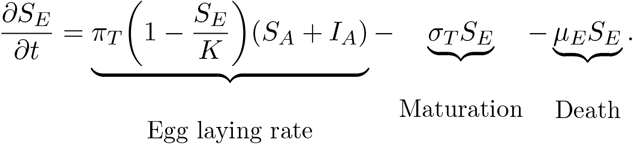

Next, we describe the dynamics of the susceptible and infectious larvae. We assume that the motion of ticks is due to their attachment to hosts they feed on [2, 34]. The rate of diffusion for juvenile ticks is denoted by *D*_*M*_. Since the diffusion only occurs when ticks are attached to hosts, the diffusion rate for juvenile ticks is the same as the diffusion rate of mice. To model the movement of the larvae in space we use the Laplacian operator Δ to denote the sum of the second partial derivatives with respect to each spatial variable, that is,

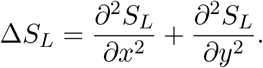

The susceptible larvae become infected after feeding on an infectious mouse at the rate *λ*_*T*_, this is the force of infection which we define as

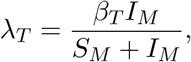

where the parameter *β*_*T*_ is the probability that a tick is infected when feeding on an infectious mouse. Both susceptible and infectious larvae mature into nymphs at the rate *τ*_*T*_, and die naturally at the rate *μ*_*L*_. Hence, the equations governing susceptible and infectious larval tick populations are given by

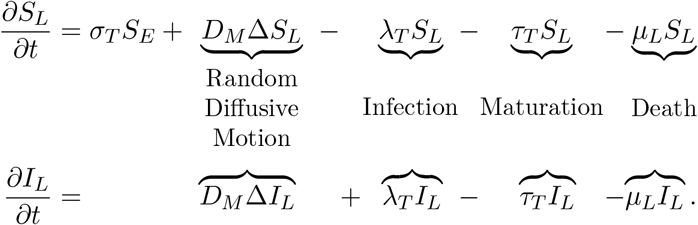

The dynamics and equations of other the other tick populations follow a similar form to the larvae and we only substitute *γ*_*T*_ for the maturation rate of nymphs to adults and *μ*_*N*_, and *μ*_*A*_ for the natural death rates of nymphs and adults, respectively, so we will not describe them in detail. Note that adult female ticks preferentially seek out deer for their final blood meal, which plays an important role in their dispersal and is the reason why adult ticks have a significantly higher rate of diffusion compared to juvenile ticks [2, 34]. We denote the diffusion rate for adult ticks as *D*_*A*_. Thus, the equations of the nymphs and adult ticks are given as

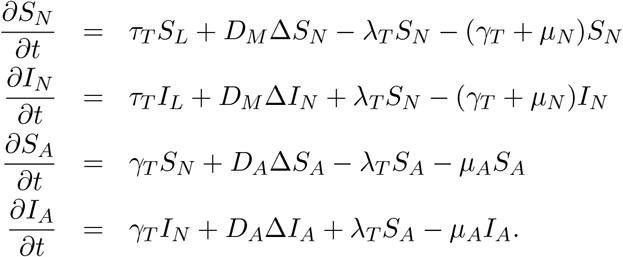

Next, we describe the dynamics and equations of the susceptible and infectious mouse populations. We assume the mouse populations increase at a constant rate *π*_*M*_ due to birth and move across the domain at the rate *D*_*M*_. The susceptible mice become infected if fed on by infectious ticks at the rate *λ*_*M*_ which is expressed as

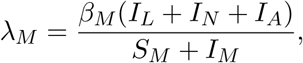

where the parameter *β*_*M*_ denotes the probability that a mouse is infected when being fed on by an infectious tick. Notice that we consider all tick life-stages (except for eggs) when calculating the force of infection since ticks at every life-stage have a tendency to feed on mice, especially if other hosts are scarce [2, 34, 35]. The mouse populations diminish due to natural death at the rate *μ*_*M*_. From this, the population dynamics for mice are described by

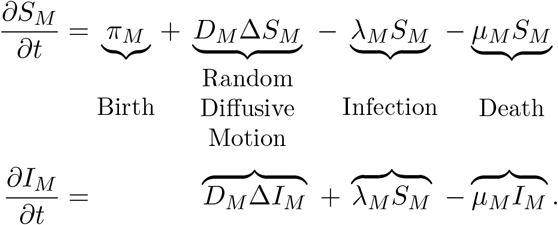

To summarize, we have the following partial differential equation (PDE) model: for *t* ≠ *nT, n* = 1, 2, …,

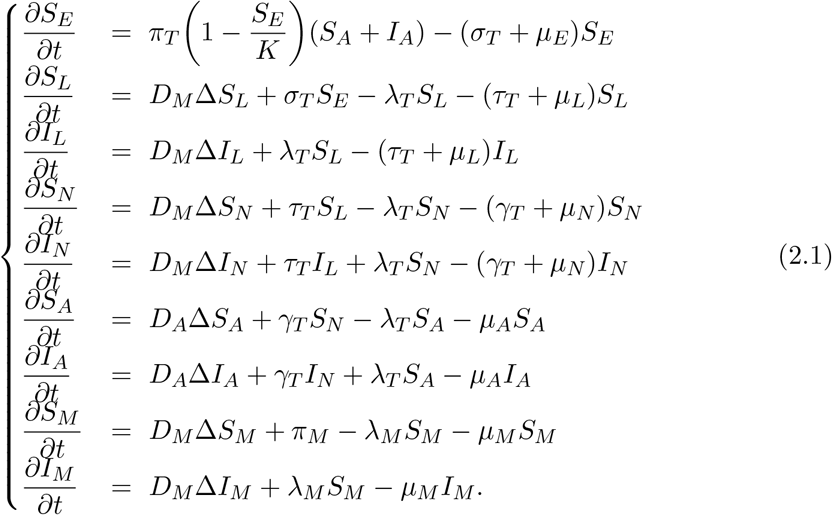

To implement prescribed fire, we assume that a certain proportion (*ν*_*E*_, *ν*_*L*_, *ν*_*N*_, *ν*_*A*_, and *ν*_*M*_) of the tick and mouse populations are reduced following a burn in a patch of the domain Ω once every *T* period of time. Thus, tick and mouse populations following a burn at time instant *t* = *nT* (*n* = 1, 2, …) are given as

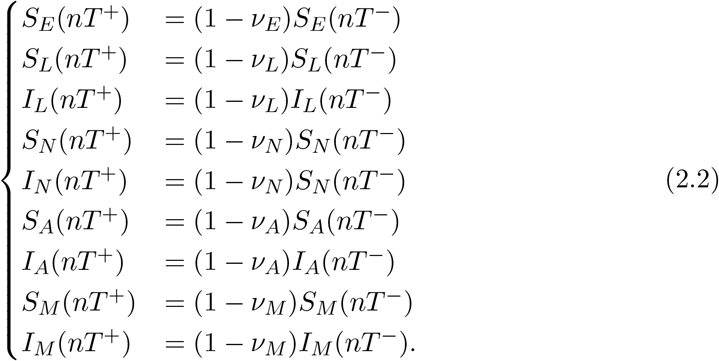

The proportion (*ν*_*E*_, *ν*_*L*_, *ν*_*N*_, *ν*_*A*_, and *ν*_*M*_) are chosen to be between zero and one; zero indicated no reduction and one indicating total loss of the tick and mice population in the burned patch. The PDE system (2.1) is subject to homogeneous Neumann boundary conditions, viz.,

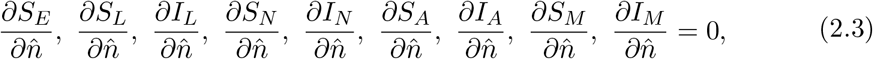

which means that no ticks are transported in or out of the domain Ω. The PDE system, the above boundary conditions, and proper initial conditions form a well-posed boundary-initial value problem.

A flow diagram for the above model is shown in Fig. 1, and variable and parameter descriptions along with the values for the parameters and references are listed in Table 1.

**Table 1:** Description of the variables and parameters for model (2.1).

| Parameter | Meaning | Value | Reference |
| --- | --- | --- | --- |
| $\pi_M$ | Birthrate of mice | 0.02 | [30] |
| $\mu_M$ | Death rate of mice | 0.01 | [30] |
| $\beta_T$ | Probability of infection in mice | 0.9 | [36] |
| $K$ | Carrying capacity for ticks | 5000 | [31] |
| $\pi_T$ | Birthrate of ticks | 456.36 | [30] |
| $\mu_E$ | Death rate of eggs | 0.0025 | [30] |
| $\mu_L$ | Death rate of larvae | 0.015 | [30] |
| $\mu_n$ | Death rate of nymphs | 0.015 | [30] |
| $\mu_A$ | Death rate of adults | 0.015 | [30] |
| $\beta_T$ | Probability of infection in ticks | 0.9 | [36] |
| $\alpha$ | Attack rate of ticks on mice | 0.002 | Assumed |
| $\sigma_T$ | Rate at which eggs develop into larvae | 0.00677 | [30] |
| $\tau_T$ | Rate at which larvae develop into nymphs | 0.00618 | [30] |
| $\gamma_T$ | Rate at which nymphs develop into adults | 0.00491 | [30] |
| $\nu_E$ | Proportion of eggs reduced following a high or low intensity burn | 1 | [26] |
| $\nu_L$ | Proportion of larvae reduced following a high or low intensity burn | 0.7421, 0.2593 | [26, 37] |
| $\nu_N$ | Proportion of nymphs reduced following a high or low intensity burn | 1, 0.7778 | [26, 37] |
| $\nu_A$ | Proportion of adults reduced following a high or low intensity burn | 0.5454, 0.9629 | [26, 37] |
| $\nu_M$ | Proportion of mice reduced following a high or low intensity burn | 0.4728 | [26] |
| $D_M$ | Diffusion coefficient for mice | 2, 0.5 | Assumed |
| $D_A$ | Diffusion coefficient for adults | 4, 1 | Assumed |

**Figure 1:**
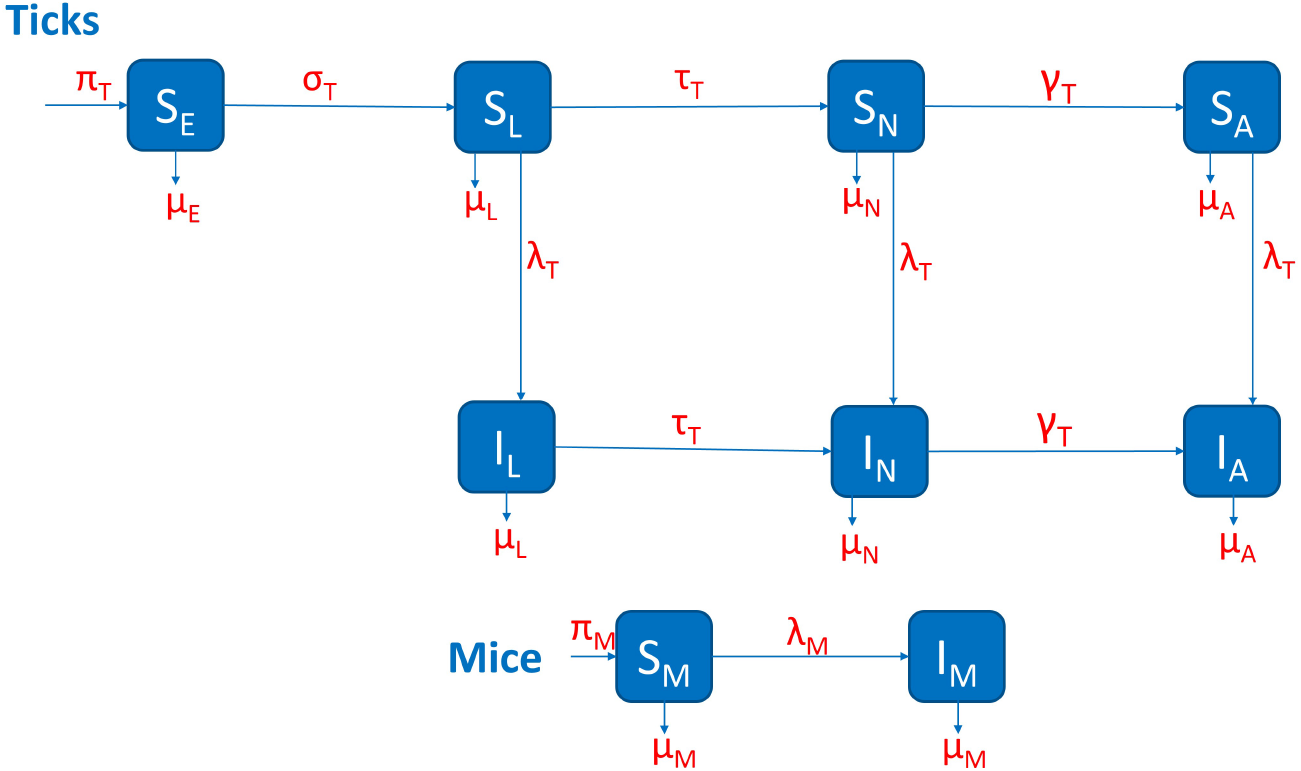
The flow diagram for model (2.1).

## 3 Numerical simulations

Our goal is to study the effects of prescribed burning in a more realistic setting compared to the ODE model presented in Guo and Agusto [29]. In the absence of diffusion, our results are the same as the results of the ODE system in [29]. Moreover, several of the results from [29] hold in a spatial setting such as the conclusions that high intensity burns are more effective than low intensity burns and that yearly burning is most effective at reducing tick populations.

First, we detail the numerical method used to solve our system of PDEs in Section 3.2. We simulate the impulsive model (2.1) and (2.2) using parameter values given in Table 1 and the proportion of ticks and mice reduced due to prescribed fire estimated in Section 3.1. However, there is little data available on the average movement of ticks. Clow et al. [38], provided an estimation of the northward range front expansion rate, but did not provide much insight into westward expansion or establishment. Adult females preferentially seek out deer for their final blood meal, which plays an important role in their dispersal and is the reason why adult ticks have a significantly higher rate of diffusion compared to juvenile ticks. In this work, we assume a rate of diffusion of 2 *km/year* for juvenile ticks and 4 *km/year* for adult ticks.

### 3.1 Estimating the proportion of ticks reduced due to prescribed fire

There is very limited data available on the survival rate of ticks following a prescribed burn, so we use what little data is available and make several assumptions in order to determine the proportions (*ν*_*E*_, *ν*_*L*_, *ν*_*N*_, *ν*_*A*_, and *ν*_*M*_). The data used to calculate the proportions for high and low intensity fires are taken from [26, 37], respectively; see Table 1. The sites for [37] were located in Hancock County, Illinois and consisted of several overstory trees including white oak and post oak trees. The burns performed during the study were low intensity since the height of the flames was largely not higher than 1 m. 54 *I. scapularis* ticks were collected over the study period and for our calculations, we assume that these were the ticks remaining following a low intensity burn. 40 larvae, 12 nymphs, and 2 adults were collected. We divide the number of ticks in each life-stage by the total number of ticks and subtract that proportion from one to get our estimate for the proportion of ticks reduced following a low intensity burn (i.e. *ν*_*L*_ = 1 − 40/54 = 0.2593). No mice or eggs were collected for this study, thus we assume that those two populations are reduced at the same rate as a high intensity fire, so our results for low intensity fires likely overestimate their effectiveness.

The sites used in the high intensity fires were located in Mendocino County, California and consisted of chaparral habitat, which is not ideal habitat for ticks, so our estimates may be higher than the true values. To calculate the proportions for high intensity fires from the data in [26], we assume that equal numbers of ticks and mice are in each site pre-burn and that any differences in the number of ticks is due to the burn performed. Then we subtract the number of ticks present in the treatment sites from the number of ticks present in the control sites and divide it by the total number of ticks in the control sites. Lastly, we subtract these values from one to give the proportion of ticks reduced following a high-intensity burn (i.e. *ν*_*L*_ = 1 − (159 − 118)/184 = 0.7421).

### 3.2 Numerical method

The system of PDEs (2.1) and (2.3) is solved using MMPDElab [39], a package written in Matlab for adaptive mesh movement and finite element computation in 1D, 2D, and 3D. The package uses the linear finite element method to discretize PDE systems in space and the implicit fifth-order Radau IIA method in time with variable time stepping. The mesh adaptivity function of the package is not used in the current work. We use a triangular mesh constructed as follows: the domain Ω is first partitioned into small rectangles of same length in each of the *x* and *y* directions. Each small rectangle is subsequently partitioned into four triangles to obtain the final triangular mesh. In our computation, we use 49 × 49 small rectangles and thus a total of 9,604 triangular elements in the mesh.

### 3.3 Global sensitivity analysis

We now perform a global sensitivity analysis (GSA) to explore which model parameters have the greatest effect on model outcomes. The results of this analysis also informs several parameter values to be used in the subsequent scenarios. Latin Hypercube Sampling (LHS) is used to create representative distributions of each of the model parameters, we generate 1000 samples for each parameter [40] to create the LHS matrix and we subsequently calculate Partial Rank Correlation Coefficients (PRCC) for each of the parameters [40]. Since there is little data available on the actual distributions of our parameters, we assume a uniform distribution for each centered on the baseline values given in Table 1 with minimums and maximums for each parameter being minus and plus 20% of the baseline, respectively.

The outcome measure here is the total number of infectious nymphs in one dimension for expediency and we perform this analysis using high-intensity burns and using low-intensity burns. We perform the analysis for a case with homogenous initial conditions as well as a case with heterogeneous initial conditions. The results are largely the same aside from slightly more significant PRCC values for diffusion of mice and adults, thus we only include the heterogeneous results here.

We also want to investigate the impact of the number of burns and the time between burns on the model outcome, so we sample those parameter values from two Poisson distributions that exclude zero. The number of burns is pulled from a Poisson distribution centered on 10 since we run the first scenario for 10 years. The time between burns is pulled from a Poisson distribution centered on 1 since a period of 1 year between burns is used for Scenario 4.1. After creating the initial distributions, any zeros in either sample are changed to ones to avoid issues with implementing the burns. These samples are combined with our LHS matrix and we subsequently calculate Partial Rank Correlation Coefficients (PRCC) for each of the parameters [40]. The results are given in Fig. 2 for low and high intensity, respectively. We also record in Fig. 3 the total number of infectious nymphs present at the end of each simulation performed from the parameter space created from the LHS method.

**Figure 2:**
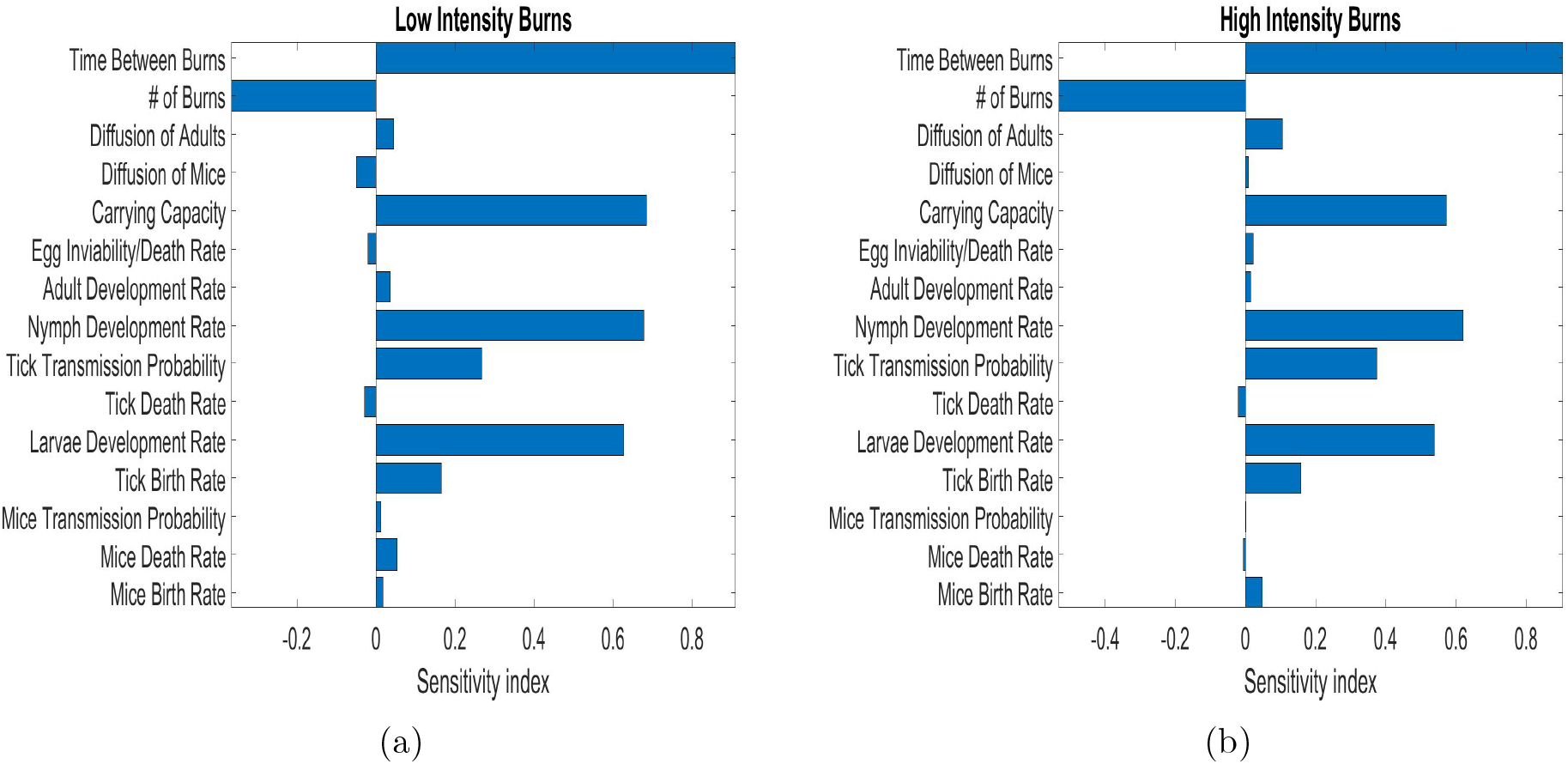
The partial rank correlation coefficients for the parameters for (a) low intensity burns and (b) high intensity burns. Values significantly greater or less than zero indicate parameters that have the greatest impact on model outcome (number of infectious nymphs).

**Figure 3:**
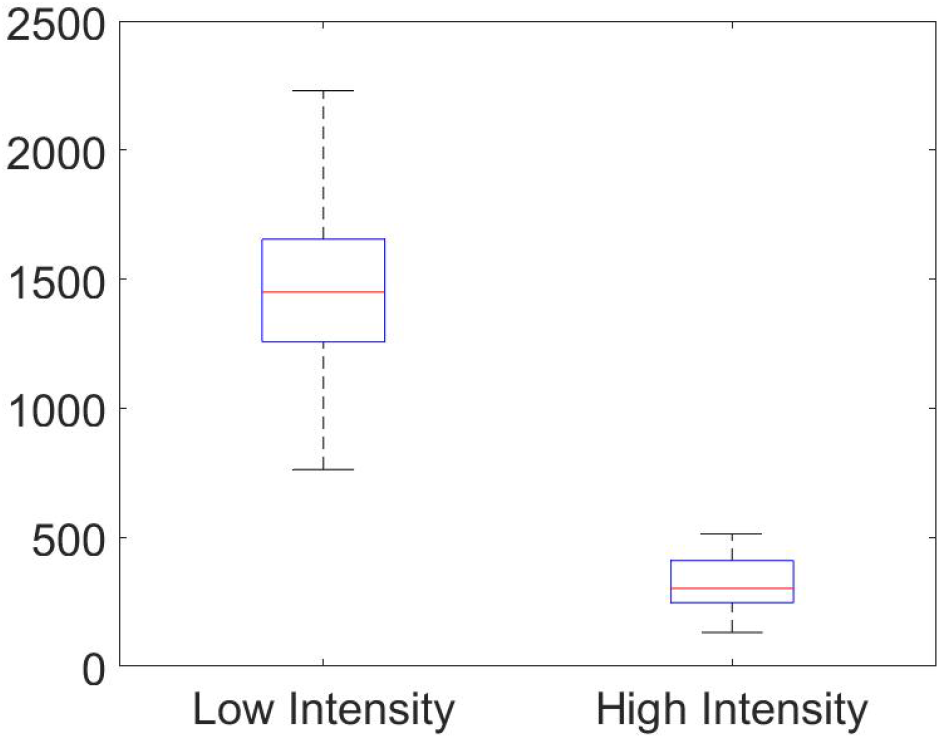
Distribution of the total number of infectious nymphs using the parameters created from the LHS. This graph displays the distribution of those totals via a boxplot. The median is shown as the red bar within the box. The upper and lower quartiles are the upper and lower portions of the box, respectively. Finally, the maximum and minimums for low and high intensity are displayed at the ends of the upper and lower dashed lines, respectively.

From the result of the sensitivity analysis in Fig. 2, we see that the significant parameters for low and high intensity burns are time between burns, number of burns, carrying capacity (*K*), nymph development rate (*γ*_*T*_), tick transmission probability (*β*_*T*_), larvae development rate (*τ*_*T*_), and tick birth rate (*π*_*T*_). Diffusion of adults is significant in the high intensity analysis, but not in the low intensity analysis.

Upon closer look at Fig. 2, we observe that the transmission probability is high with high intensity burns which will favor more infections, but the high intensity environment is hostile since vital parameters like tick developmental rates, and the carrying capacity have a relatively low impact on the number of infectious nymphs, unlike the low intensity burn domain which has high sensitivity indices for these parameters.

Looking at the time between burns, and number of burns closely, we see that regardless of whether we are performing low or high intensity burns, the timing between burns remains crucial, indicated by a PRCC value near one in both cases. This means that even if there is a case where low intensity burning is the only type possible for any reason, whether that be due to lack of fuel or proximity to homes or businesses, there is a significant negative effect on the total number of infectious nymphs present in that habitat as time between burns decreases. This is reflected in the results by the large positive PRCC value for time between burns. This positive value indicates a positively correlated relationship between this parameter and the model outcome, so when the value of this parameter increases, so does the number of infectious nymphs and visa versa.

We also observe that the number of burns being performed over time plays an important role, but less so than timing between burns. The number of burns plays a slightly larger role in the high intensity case since many more nymphs are killed off with each burn compared to the low intensity case (See Fig. 3). The negative PRCC value for this parameter indicates that it has a negatively correlated relationship with the model outcome, so when this parameter increases, the number of infectious nymphs tends to decrease.

Considering tick and host diffusion across the domain, we observe that diffusion of the ticks and hosts does not have significant effects on the model outcome. It may be that we have not tested a wide enough range for these parameters to see their effect.

All these observations help inform some model parameters for the scenarios presented in the following sections, namely, the use of high intensity burns, and a reasonable time between burns of one year, unless otherwise stated. All other parameters are kept at their baseline values for these scenarios unless otherwise stated.

Now using the parameters created from the LHS, we see in Fig. 3 as with the results from [29] that in a spatial setting high intensity burns are more effective than low intensity burns at reducing tick populations. Furthermore, we see that the minimum of the boxplot for low intensity burns is more than the maximum from the high intensity burns.

## 4 Prescribed fire scenarios

In this section, we examine the effect of prescribed burns and diffusion in different scenarios that may arise in various environments in the real world.

### 4.1 Scenario 1: Prescribed fire in a homogeneous domain

The effects of consistent burning and diffusion are addressed in this scenario where ticks are distributed evenly across the domain Ω of size 30 *km* × 30 *km* and a prescribed burn occurs in a 10*km*×10*km* block in the middle of the domain every twelve months for ten years. This equates to roughly 2500 acres of burned area, which is a large prescribed fire typical in larger uniform environments such as oak woodlands or grasslands [41]. We assume an initial condition of 500 infectious nymphs at every point in the domain, meaning that Lyme disease is already endemic in this area. Fig. 4 shows the simulation results for this scenario.

**Figure 4:**
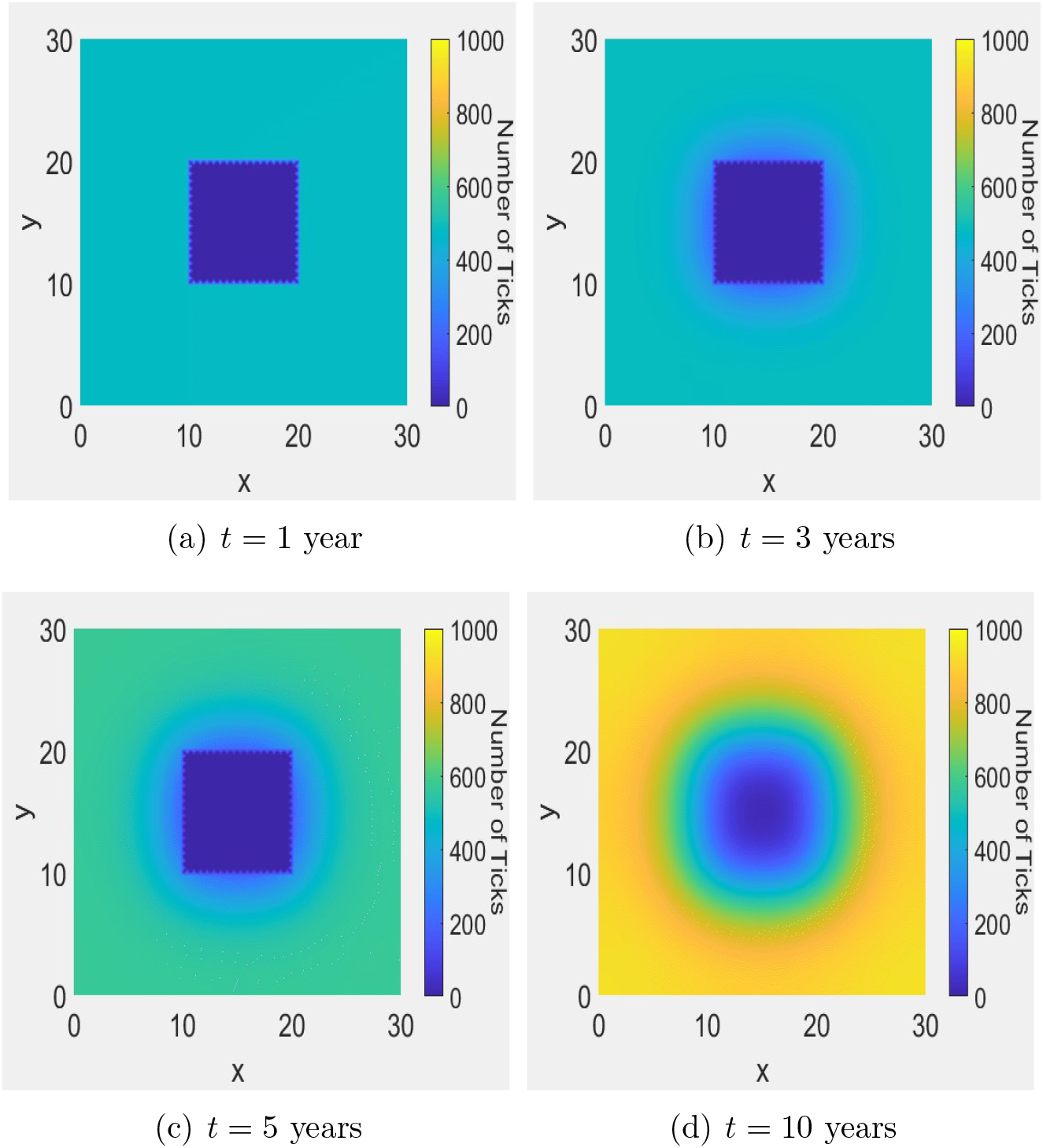
The distribution of infectious nymphs in a homogeneous domain at various times.

From these results, one can see an immediate reduction in the number of ticks at *t* = 1 following the burn in Fig. 4(a); unfortunately, this is followed by ticks diffusing into the burned area and a slow recovery of the burned patch in terms of the number of ticks. At *t* = 3 (Fig. 4(b)), we can see just how effective burning is at reducing the number of ticks in the burn site and the effects of diffusion begin to become clear. Ticks begin to diffuse into the burned environment during the year following a burn and increase the recovery rate of ticks in that area while simultaneously decreasing the number of ticks in the area surrounding the burn. The recovery rate of ticks is still low enough that the number of ticks remains lower in the consistently burned area even after long periods of time (see Fig. 4(c)). By the end of the ninth year, just before a burn in the tenth year, we can see in Fig. 4(d) that the number of ticks in and around the burn site has been reduced significantly compared to the areas near the edge of the domain that were unaffected by both the burns and the diffusion of ticks into the burned area.

To further explore the effectiveness of prescribed burns in a homogeneous domain, we examine the effect of a different number of burns and time between burns over a period of 10 years.

#### Effects of a different number of burns and time between burns

In Fig. 5(a), we examine the effect of the number of burns. In other words, we perform a set number of yearly burns and let the simulation run for a total of 10 years to determine whether or not the number of burns has a significant impact on the number of infectious nymphs present in the burned area at the end of the simulation. For example, the case with 4 burns means that 4 yearly burns are performed starting at *t* = 1 and no burns are performed for the rest of the years. The results indicate that consistent prescribed burning is an effective control method for tick populations. In addition, we see that there is nearly a linear relationship between the number of burns performed and the percentage of ticks that remain at the end of the scenario, so the more burns that are able to be performed, the better.

**Figure 5:**
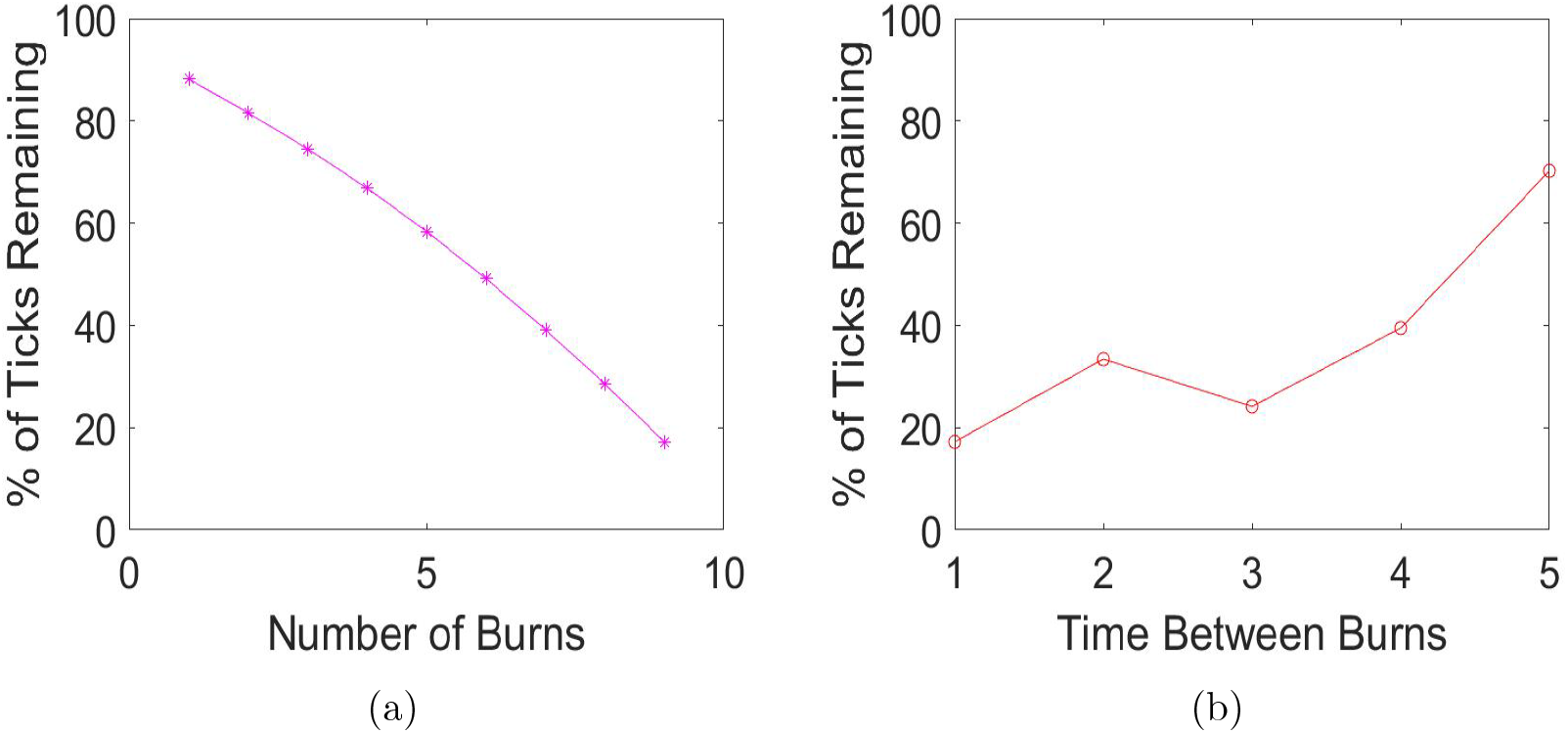
Simulation of model (2.1) for different number of burns and time between burns. (a) The number of burns performed yearly starting at *t* = 1 vs the percentage of ticks that remain after a 10 year simulation. (b) The number of the years between burns vs the percentage of ticks that remain after a 10 year simulation.

Next, in Fig. 5(b) we look to the time between burns to explore the effectiveness of burns at different times since yearly burning might not be possible in every environment. Note that the burning pattern in this situation is different from that considered in Fig. 5(a). For example, with the number of the years between burns being 4 the burn is performed every four years with the first burn occurring at *t* = 4 and the total number of burns is 2. We observe that the best case is a yearly burn, this of course agrees with our earlier result from the sensitivity analysis. Moreover, we see that burning at any interval is very effective at reducing the number of ticks in an area where they are endemic. Even a single burn every five years led to a 30% reduction in the number of ticks compared to the unburned case.

#### Effects of different burn patch sizes

Finally, we explore the effect of changing the patch size of the burn being performed. To better compare the relative effectiveness of each size of the burn patch, we divide the number of infectious nymphs remaining in the burned area at the end of a simulation by the number of infectious nymphs present in a patch of the same size in a simulation that ran for ten years, but had no burning occurring. This measure allows us to compare the effectiveness of burning with larger and smaller patches. We see in Fig. 6 that the bigger the patch, the more effective burning is at reducing the number of infectious nymphs.

**Figure 6:**
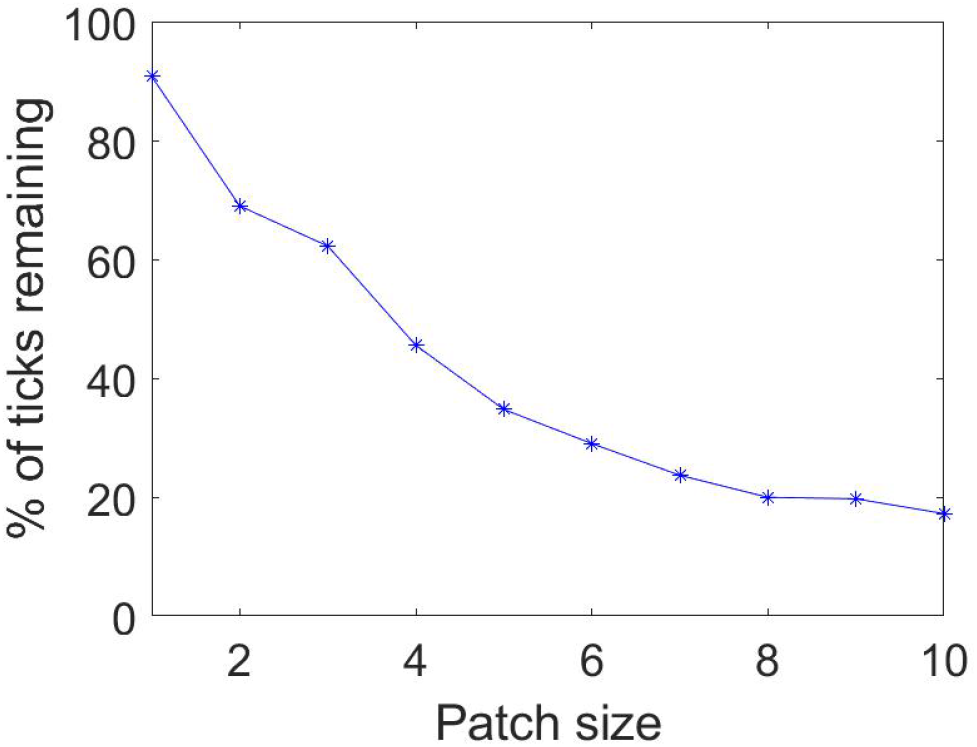
Scenario 1: The percentage of the number of infectious nymphs remaining in the burned area after 10 years of yearly, high intensity burns is plotted against the patch size.

Thus, patch size plays a very important role in determining the effectiveness of a burn. One may argue that this is due to the simple fact that we are killing off more ticks as burn size increases, but notice that the percentage remaining decreases non-linearly as the patch size increases. This can be attributed, in part, to the effect of diffusion. Since nymphs diffuse at a rate of 2 *km/yr*, we expect that a burn patch whose length is less than that rate will be significantly less effective than one that is greater than that length and this is reflected in our results based on the large increase in effectiveness, which is signified by a large decrease in the percentage of ticks remaining, when moving from a patch size of 1 *km* × 1 *km* to a patch size of 2 *km* × 2 *km*. We see a similar reduction when moving from a patch size of 3 *km* × 3 *km* to 4 *km* × 4 *km*. This is due to the effect of adult tick diffusion, which occurs at a rate of 4 *km/yr*. There are diminishing returns as patch size increases, but if the goal of a burn is reducing the prevalence of ticks, then the larger the patch, the better. The relationship seen here highlights the need for additional studies on the aggregate movement of ticks in the Midwest as this data can inform burning management strategies.

In the next section, we further explore the effectiveness of prescribed burns in a heterogeneous domain that captures different environments that ticks can be found.

### 4.2 Scenario 2: Prescribed fire in a heterogeneous domain

In order to explore how burning in different environments might affect tick populations, Scenario 2 has a nonuniform domain with *x* > 15 *km* being an wooded environment for ticks, which typically leads to better survival rates and easier traversal. This is interpreted as the ticks having a higher rate of diffusion compared to *x* < 15 *km*, which is considered grassland with burns occurring in 10 *km* × 10 *km* blocks on opposite corners of the domain. Grassland is a less suitable environment for ticks because it generally has higher temperature and lower humidity relative to a wooded environment, which also tends to have significant leaf litter so ticks have plenty of space to hide when temperature increases or humidity drops. Ticks in the wooded environment have a rate of diffusion 4 times greater than those in the grassland environment. The relatively low rates of diffusion in both environments means that there is no significant effect of the ticks in the wooded environment on the area that is burned in the grassland environment and visa versa.

Fig. 7 shows the results of Scenario 2. By the time of the first burn at *t* = 1 (Fig. 7(a)), we can see that the results are very similar for the two environments. However, by *t* = 5 (Fig. 7(c)), there is a clear difference between them. Since the rate of diffusion is higher for *x* > 15 *km*, the number of ticks surrounding the area that has been burned is lower compared to the area around the burn for *x* < 15 *km*. At the end of the final year, the effects of burning in a wooded environment indicate that it is more effective in reducing the number of ticks when considering the total area in and around the area that is being burned. To verify this, we calculated the number of ticks in each area that is burned as well as the number of ticks present in the regions *x* < 15, 15 < *y* < 30 *km* and *x* > 15, *y* < 15 *km* (i.e. the upper left and lower right quadrants, respectively). Interestingly, the number of ticks present in the grassland burned area was less than the woodland burned area, however the number of ticks present in the upper left quadrant was greater than the number of ticks present in the lower right quadrant. This is compelling evidence that burning in an environment that is better suited to ticks is significantly more effective at reducing the number of ticks both in and around the area that is burned. As time goes on and burning continues, this negative effect on ticks becomes more and more noticeable. On the other hand, if the goal of the burn is strictly to reduce the number of ticks in the area that is burned, then burning in the grassland area would lead to a better outcome. This scenario indicates that careful consideration should be taken when planning prescribed burns as the goal of the burning will inform the best areas for that burning to occur.

**Figure 7:**
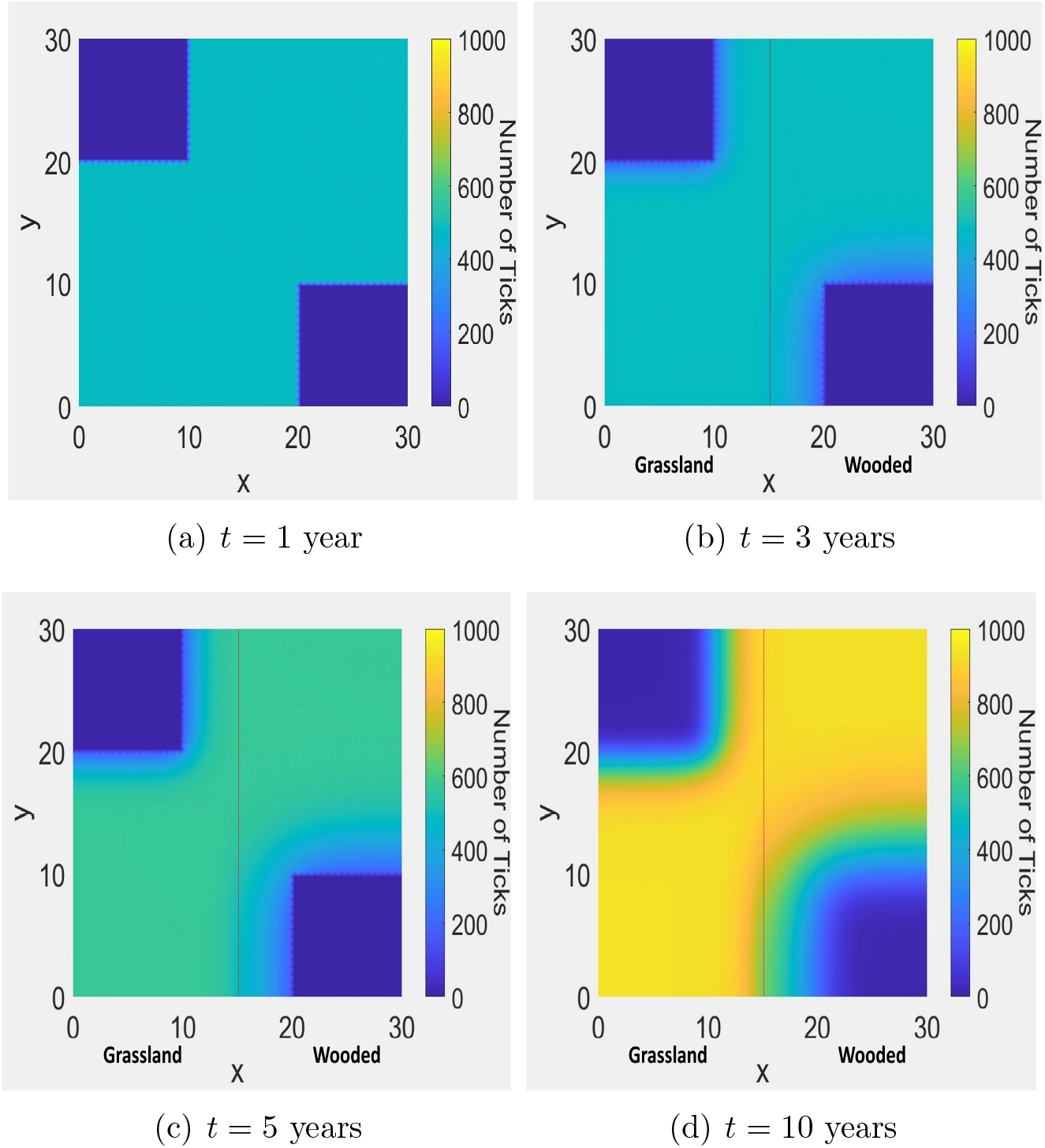
The distribution of infectious nymphs in a heterogeneous domain at various times.

### 4.3 Scenario 3: Prescribed fire at the invasion front

In this final scenario, we explore the effect that prescribed burns have on the invasion of ticks into a new area. Suppose that ticks are discovered in high numbers in the region *x, y* < 5 *km* and burns are performed for *x, y* < 10 *km* in an attempt to halt the spread of ticks into the rest of the domain. Note that susceptible hosts are assumed to be present throughout the domain, otherwise establishment would not be possible. These final scenarios (Figs. 8-9) contain one example without burns and three different examples with burning in order to explore the applicability of fire in reducing the number of ticks invading into a new area. For the latter three variations, we only show the results at the end of the simulation (that is, at *t* = 20).

**Figure 8:**
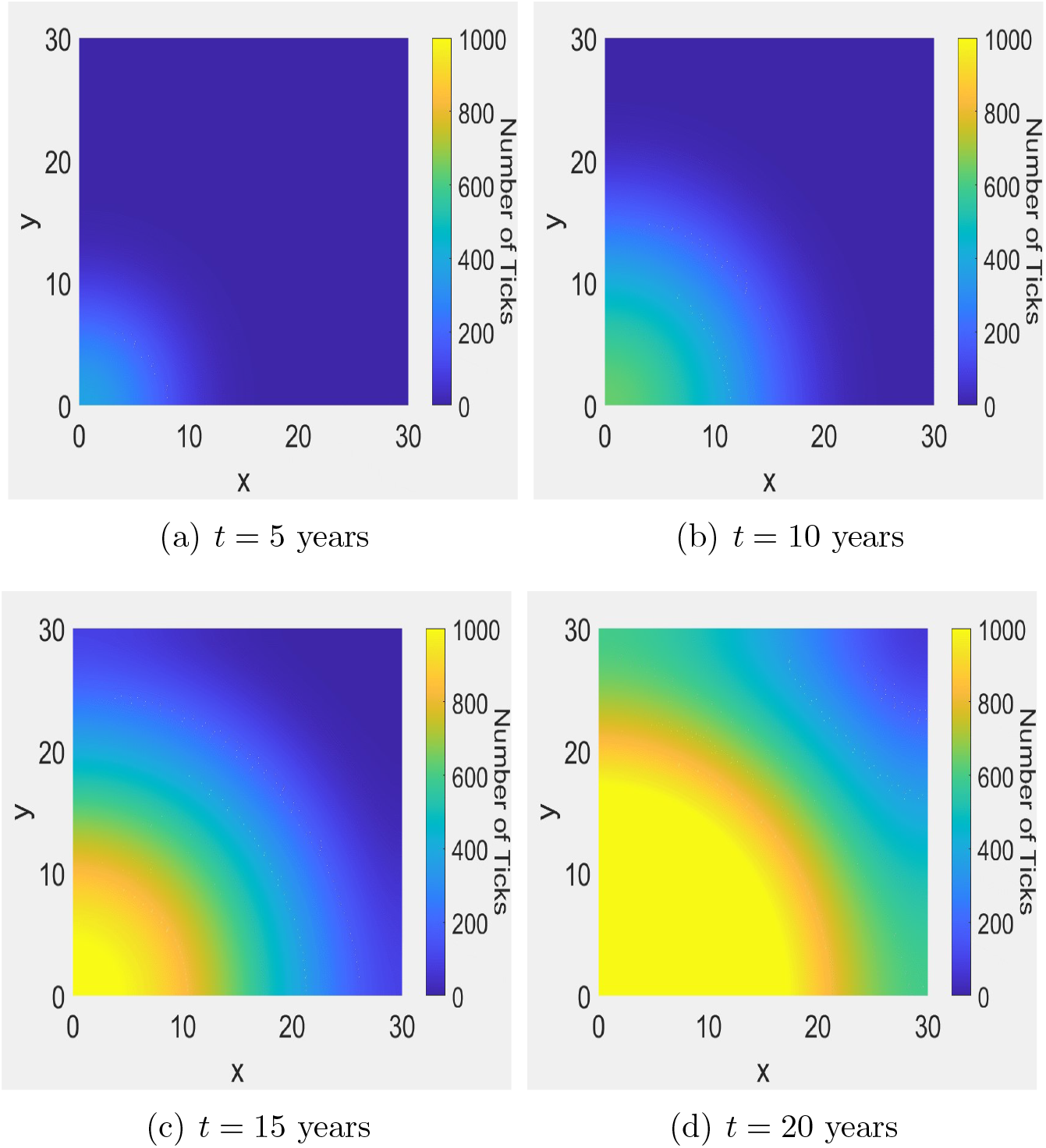
The distribution of infectious nymphs for Scenario 3 without prescribed fire at various times.

**Figure 9:**
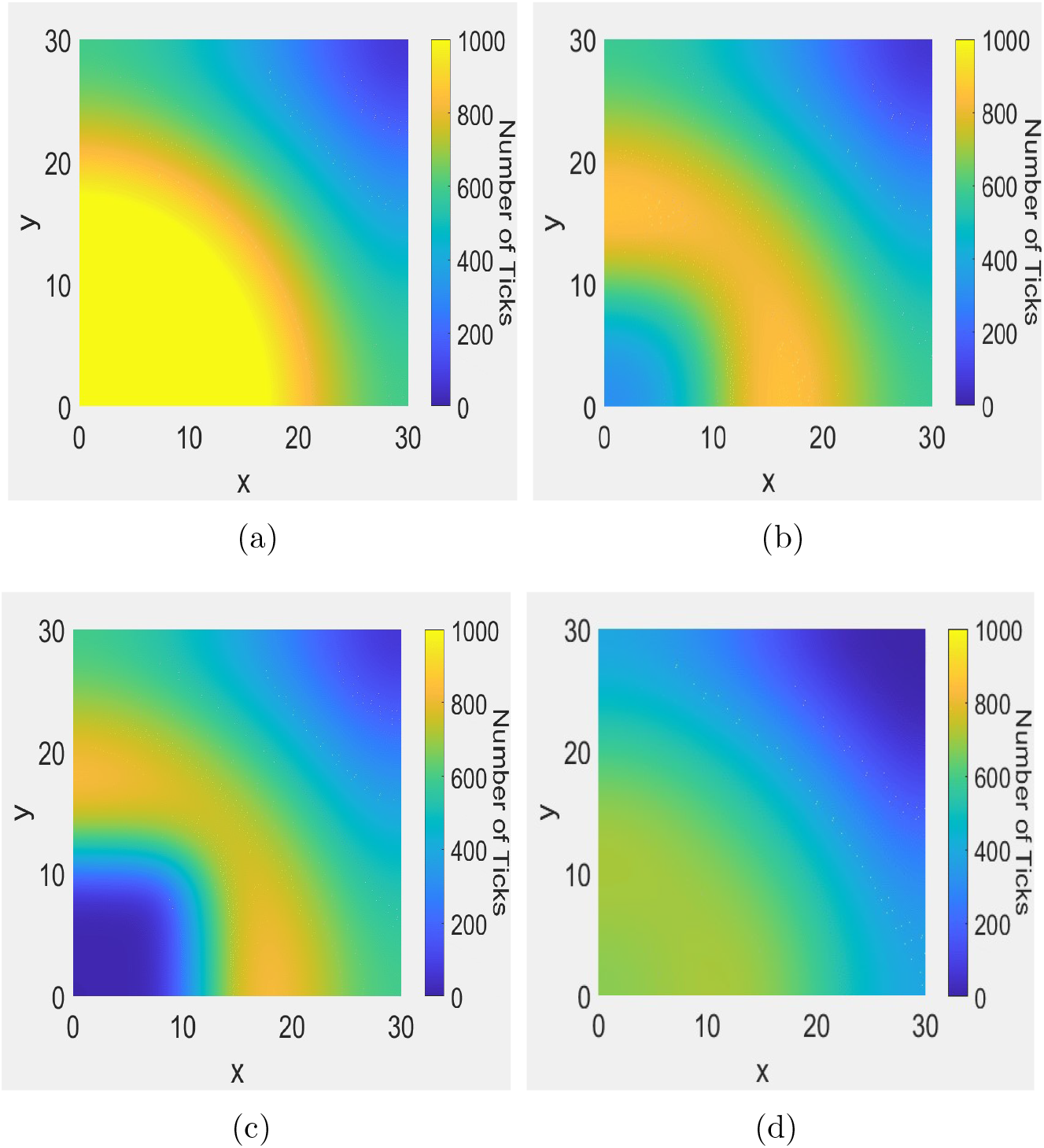
The distributions of infectious nymphs at *t* = 20 years for Scenario 3 with different burning patterns. (a) No burns. (b) Burning implemented every 5 years. (c) 10 years of no burning followed by yearly burning for the remaining decade. (d) Yearly burning implemented for the first 10 years and no burns later.

In this last section, we further explore the effectiveness of prescribed burns at the invasion front where ticks are establishing in a new environment.

#### Invasion front without prescribed fire

Figs. 8(a) - 8(d) show what occurs after 20 years of uninhibited spread. A traveling wave is formed and begins to propagate across the domain. This situation is likely occurring in many places across the Midwest and further North as ticks continue to spread across the continent.

When left unchecked, it is clear why tick numbers have been increasing over the past several decades. The lack of fire management in many areas combined with the effects of climate change and an abundance of hosts have made many areas of the United States prime environments for ticks and once they are established, they can spread rapidly. Next, we analyze the results of the three different burning examples mentioned previously.

#### Invasion front with prescribed fire

The first burning regime is one in which prescribed burns are implemented every 5 years after ticks are discovered in the corner of the domain. Fig. 9(b) displays the results of the simulation after 20 years. We see that burning every 5 years has a significant impact on the number of ticks remaining at *t* = 20 years. Compared to the example without burning, there is a 23% decrease in the number of ticks remaining. However, we can also see that burning every 5 years has had almost no effect on the spread of ticks into the region. The edge of the wave is in virtually the same position as the case without burning.

The second burning example, which is depicted in Fig. 9(c), is a case where the high prevalence of ticks in the corner of the domain is not noticed until the tenth year of the simulation and a yearly burning regiment is performed for a decade. In this case, it is clear that even after a decade of burning, burning alone is not entirely effective at preventing the spread of ticks. There has been a significant reduction in the percentage of ticks, with 32% of ticks being killed off compared to the unburned case.

The final case is depicted in Fig. 9(d). This is a case where the invading ticks are detected early in the first year and a yearly burning regiment is planned and executed for one decade. Again, we let this example run until *t* = 20 years to evaluate the effectiveness of the approach compared to the previous examples. By *t* = 20, we can see that despite a decade of high intensity burning, some of the ticks have survived and made their way to a majority of the rest of the domain. This indicates that if ticks are noticed early enough, burns may be able to limit the number of ticks that are able to spread into a new area, though establishment may be inevitable. Pairing prescribed burns with other forms of tick population management would likely slow and possibly prevent spread and establishment. Comparing this with the unburned example, we have a 42% reduction in the number of ticks at the end of the simulation. From this, we can conclude that burning as early as possible is crucial to the effectivness of prescribed burns on the invasion of ticks. The results of this final scenario paint a difficult picture. Many areas in the Midwestern United States are experiencing or expect to experience increasing numbers of ticks as the effects of climate change worsen. If prescribed burning is to be used as an effective control method, then the time for action is now as waiting any amount of time gives these organisms the chance that they need to establish themselves firmly enough that prescribed fire can only be used as a mitigation strategy rather than as part of a strategy for eradication.

## 5 Discussion

As stated previously, one of the primary papers in this study is Guo and Agusto [29]. This work on the effects of prescribed fire on tick populations provided crucial information that guided several of the numerical simulations considered, namely, those in Scenario 1. Since one of the main conclusions of the paper is that high intensity fires are significantly more effective than low intensity fires, we do not need to explore the two variations too deeply to come to a similar conclusion. Thus, we performed an extension of the sensitivity analysis given there and further analyzed the effects of those results in Scenario 1. Based on the results from Figs. 2 and 3, combined with the results from [29], we conclude that further testing of the effects of low intensity burning, while interesting, has little practical importance since low intensity fires are unable to reduce the number of ticks in an area at a similar rate as high intensity fires, so high intensity fires should be used whenever possible.

Our results have several implications for the future of prescribed fire as a method of control for tick populations. In particular, Scenario 1 highlights the waning effectiveness of burning as patch size decreases, which may explain conflicting results observed by several studies conducted on the topic thus far [13, 14, 15, 17, 20, 22, 24, 28]. As stated above, many of these studies focused on small plots of land and were sometimes performed on previously unburned sites. Burning on small plots can have little to no effect depending on the recovery rate of ticks which is governed by several factors including the average movement of hosts in the region as well as the environment in which the burn is being performed. As the average rate of movement of hosts in and around the area that is being burned increases, an example of this being the transition time between winter and spring when both small and large mammalian hosts become more active, a larger and larger area is required for a prescribed burn to significantly reduce the number of ticks in that area. Burning previously unburned plots of land has increasing effectiveness as subsequent burns are performed, and if patch size is large enough and intensity is high enough, there can be a significant reduction in the number of ticks in the burned area. Since the site was previously unburned however, recovery of ticks can occur relatively quickly since a single burn does little to reduce the number of ticks around the area being burned, thus multiple burns should be performed over longer periods of time to consistently reduce the number of ticks in and around the area being burned.

Based on our results, we theorize that the effectiveness of prescribed burning is dependent on the average movement of hosts in and around the area being burned. The study sites used in [12, 19] combined with our results from Scenario 2 paint an interesting picture. The plots used in [12, 19] mostly consisted of pine and mixed-pine forested environments and Georgia and Florida have large deer populations and few white-footed mice. The lack of this extremely viable host should further inhibit recovery of ticks following a burn and that seems to hold true based on the observed results. Since there was also a reduction in the prevalence of certain diseases in the population [19] may indicate that a combination of hosts are required for effective tick and disease reestablishment or that host composition is extremely important when determining the effectiveness of a prescribed burn. Thus, several factors need to be considered when deciding on the best control methods for ticks in an area, including the type of tick hosts present in and around the area and their prevalence, the local environment, and the type of burn (low vs. high) that is able to be performed regularly.

When we look at all of this information alongside the results presented in Scenario 3, there is major concern for several areas in the United States. Those states located in the Great Plains seem to be particularly vulnerable to invasion of ticks. This is due to the various factors mentioned in the previous paragraph. Since the area is largely covered by grassland and farmland and these environments are either not suited for consistent burning or more quickly provide renewed cover for ticks via fast-growing grasses, prescribed burning as a means of tick population management in these areas may not be as feasible or effective. This is especially true considering the limited effect of burning on tick establishment in our simulations.

## 6 Conclusions

In this study we extended the model provided in Guo and Agusto [29] in order to explore how the effects of prescribed burning impact tick population dynamics in a spatial setting. The results of this study indicate that prescribed fire is an extremely useful tool for tick population management, but a limited tool for prevention of tick establishment. The relationship between burn patch size and percentage of ticks remaining in the burned area was explored and this relationship coupled with other results presented here provide the insight that the effectiveness of prescribed burns can vary widely depending on biological factors related to the aggregate movement of ticks and their hosts. When the patch being burned is less than the rate of diffusion of ticks, we see a less significant effect of burning compared to larger patch sizes which yield a much lower proportion of ticks remaining after a decade of regimented burning. The results from Scenario 2 support the idea that the location of burns is crucial depending on the goal of the burning. This is due to the fact that burning in areas where ticks cannot easily spread due to increased rates of desiccation or lack of hosts, such as grasslands which tend to be hotter and drier than forested environments, is only effective at reducing the number of ticks in the area the that is burned, while burning in a more advantageous environment leads to a greater overall reduction in the number of ticks over time both in and around the area that is burned. Since the location of burns plays a crucial role in determining the effectiveness of prescribed fire, not all areas that ticks inhabit will be suitable for this method of control. In those cases, other methods of control mentioned in the introduction may be more effective over time. Lastly, Scenario 3 indicates that so long as hosts are available in an area where ticks are not yet present, ticks will spread and prescribed fire may need to be used in conjunction with other control methods for eradication to have a chance of success. It is our hope that this paper informs those that perform controlled burns in the future on how and where those burns should be executed for the purpose of tick population management.

There are several future directions that are being considered. First and foremost, incorporation of seasonality into the model that was used is crucial to ensure that our results hold when considering a more realistic situation regarding tick population dynamics. It is likely that if a burn is performed during periods of extremely low tick activity, then it would be considerably less effective, thus timing of burns must be explored once this additional layer of complexity is added to the current model. A more subtle question in this vein is regarding the effectiveness of prescribed burns during peak activity of the different life stages of ticks. Is it more effective to perform a burn when tick nymphs are more active, or when larvae are most active, or adults? This is a very interesting question and one that has no clear answer since each life stage handles environmental changes differently. A more realistic incorporation of prescribed fire would also be beneficial in finding out the level of detail needed to best model the effects of prescribed fire. For example, [12] indicates that prescribed burning changes the micro-climate in forested habitats, making it hotter and drier over time. These changes are important to consider since slight changes in temperature and humidity can be devastating to the survival rate of ticks. The changes would likely be incorporated via a micro-climate function that contains all of the relevant information for calculating temperature and humidity over time and space including how these are affected by prescribed burning. These issues will be addressed in future publications.

## Data Availability

All data produced in the present work are contained in the manuscript

## Acknowledgement

This research was supported in part by National Science Foundation under EPSCOR Track 2 grant number 192094.

